# Constructing a Finer-Grained Representation of Clinical Trial Results from ClinicalTrials.gov

**DOI:** 10.1101/2023.10.25.23297572

**Authors:** Xuanyu Shi, Jian Du

## Abstract

Randomized controlled trials are essential for evaluating clinical interventions. ClinicalTrials.gov serves as a primary repository for such data, yet extracting and synthesizing information from it remains challenging. This study introduces a novel methodology for constructing a detailed arm-centered representation of clinical trial results, moving beyond the traditional PICO (Patient, Intervention, Comparison, Outcome) framework. The representation attentively uncovers both efficacy outcomes and adverse drug events in safety outcomes, promoting a dual-faceted understanding of intervention effects. Through a structured acquisition, extraction, and initialization process, we present a knowledge graph incorporating arm-level efficacy with safety results, categorizing outcomes into three distinct groups: biomarkers, patient-reported outcomes, and clinical endpoints. The goal is to bridge the gap between the generally described searchable design information and the specifically detailed reported results. This approach aims to offer a structured dataset towards better utilization and interpretation of ClinicalTrials.gov data, facilitating a more feasible and complete evidence synthesis practice to include both positive and negative results hidden in clinical trials registries.

## Background & Summary

Clinical trial, specifically randomized controlled trial (RCT), is a method of perspective, random allocated double-blinded clinical study. The goal of carrying out a clinical trial is to evaluate the efficacy and safety or a single or multiple clinical interventions. It is also a requisite step before a drug is approved for market release and sale. In the new evidence-based medicine pyramid, randomized controlled trials are rated as the highest position, representing the most reliable medical evidence^1^. In today’s medical research landscape, the accuracy and accessibility of clinical trial data are paramount to ensuring research quality and advancing scientific progress. ClinicalTrials.gov (CT.gov) serves as a pivotal repository for such data, hosting valuable information concerning numerous clinical trials. However, extracting and synthesizing this information for further analysis remains a challenging endeavour.

The unique value inherent in CT.gov data has been demonstrated through various comparisons and analyses. When compared with results from PubMed, it has been noted that CT.gov often contains a more comprehensive report of adverse events^2^. In CT.gov, safety results were reported at a similar rate as in peer-reviewed literatures, with more thorough reports of certain safety events^3^.

Discrepancies between different reports further underscore the necessity for a more systematic method of information retrieval from CT.gov. Selective reporting, as discussed in JAMA^4^, illustrates the pressing need for utilizing CT.gov as a tool for evidence synthesis. Although calls for leveraging CT.gov have been made, including in papers published by BMJ^5^, the absence of automated tools for such a task is conspicuous. Despite its seemingly structured facade, CT.gov presents substantial usability challenges which hinder the extraction of published results.

Treating CT.gov research findings as a dataset embodies the principle of “process once, use many times,” transforming these findings into a computable body of knowledge. This approach stands to significantly aid evidence synthesis. Despite a study from Johns Hopkins University suggesting that CT.gov has not altered the conclusions of systematic reviews^6^, we argue that the limited sample size in their analysis may have influenced these findings. Results of randomized controlled trials (clinical trials, or RCT), are either published in scientific articles, or reported on registry platforms, such as ClinicalTrials.gov (CT.gov), International Clinical Trials Registry Platform (ICTRP), and Chinese Clinical Trial Registry, ChiCTR). Researchers found that in the 91 trials with reported results on ClinicalTrials.gov and published in high-impact journals, only 52% primary efficacy end points were described in both sources and reported concordant results^7^. One of the possible and biggest explanation for this phenomenon is selective publication or publication bias of clinical trials, which means there exists a higher reporting rate of positive results in published literature compared to that in registries^8^.

This study intends to construct a finer-grained representation for arm-centered clinical trial results by integrating efficacy and safety information. Our methodology enables: 1) a detailed representation of “intervention” entities, transcending the traditional PICO (Patient, Intervention, Comparison, Outcome) concept representation; 2) a systematic unveiling of positive and negative results in efficacy outcomes; 3) a systematic disclosure of adverse drug events (ADE) in safety outcomes; 4) a dual-faceted understanding of intervention effects from both efficacy and safety perspectives. 5) By providing a structured, ready-to-use dataset, we aspire to offer a new data source for meta-analysis, thereby facilitating a more discoverable dataset to enhance evidence-based health decision-making.

The discrepancy between PICO information in the design phase and reported results underscores the necessity for real meta-analysis which mandates results-oriented information. Our study seeks to bridge this gap, fostering a more thorough and reliable synthesis of evidence that can potentially elevate the standards of medical research and practice.

### Existing studies

There have been studies focusing on the structuring trial results data. MedicineMaps ^9^ introduced a schema that represents clinical trial results from literature. The schema lost information about the comparators, and was annotated manually. EvidenceMap^10^ extracted PICO with observation data and efficacy relationships from 80 COVID-19 clinical trial abstracts. Similarly, entities such as ‘two groups’ were also labeled as interventional, losing the ability to build comparable relationships. TrialStreamer^11^ and TrialSummarizer^12^ are tools that extracts results from clinical trials in a large scale. For data from registry platform, CTKG^13^ is a large knowledge graph version with embedding analysis of studies in the ClinicalTrials.gov database. The knowledge graph follows the basic structure of ClinicalTrials.gov, with nodes being sections and edges being ‘has’. In ‘outcome_analysis’, same as the representation in ClinicalTrials.gov, the compared arm groups were not separated as intervention versus comparator as well.

### In this study

We introduce an arm-level representation for efficacy and safety results in studies registered on ClinicalTrials.gov. We optimize the data structure of clinical trials and present study results with one-to-one comparable relationships. Each efficacy result is represented by a relationship from an intervention arm group to the outcome, with the efficacy as the relationship value, the comparator arm as an attribute. Each safety result is represented by a relationship from an arm group to an adverse event, with the number of affected subjects as an attribute.

To represent arm-level efficacy and safety results in a knowledge graph, we started with acquiring registered clinical trials from register platform clinicaltrials.gov, followed by extraction of statistical significance and adverse events from reported results. After standardizing arm groups and outcomes, we eventually constructed a knowledge graph including results nodes, relationships and related attributes. Fig. 1 shows the overall workflow of this study.

**Figure 1.**
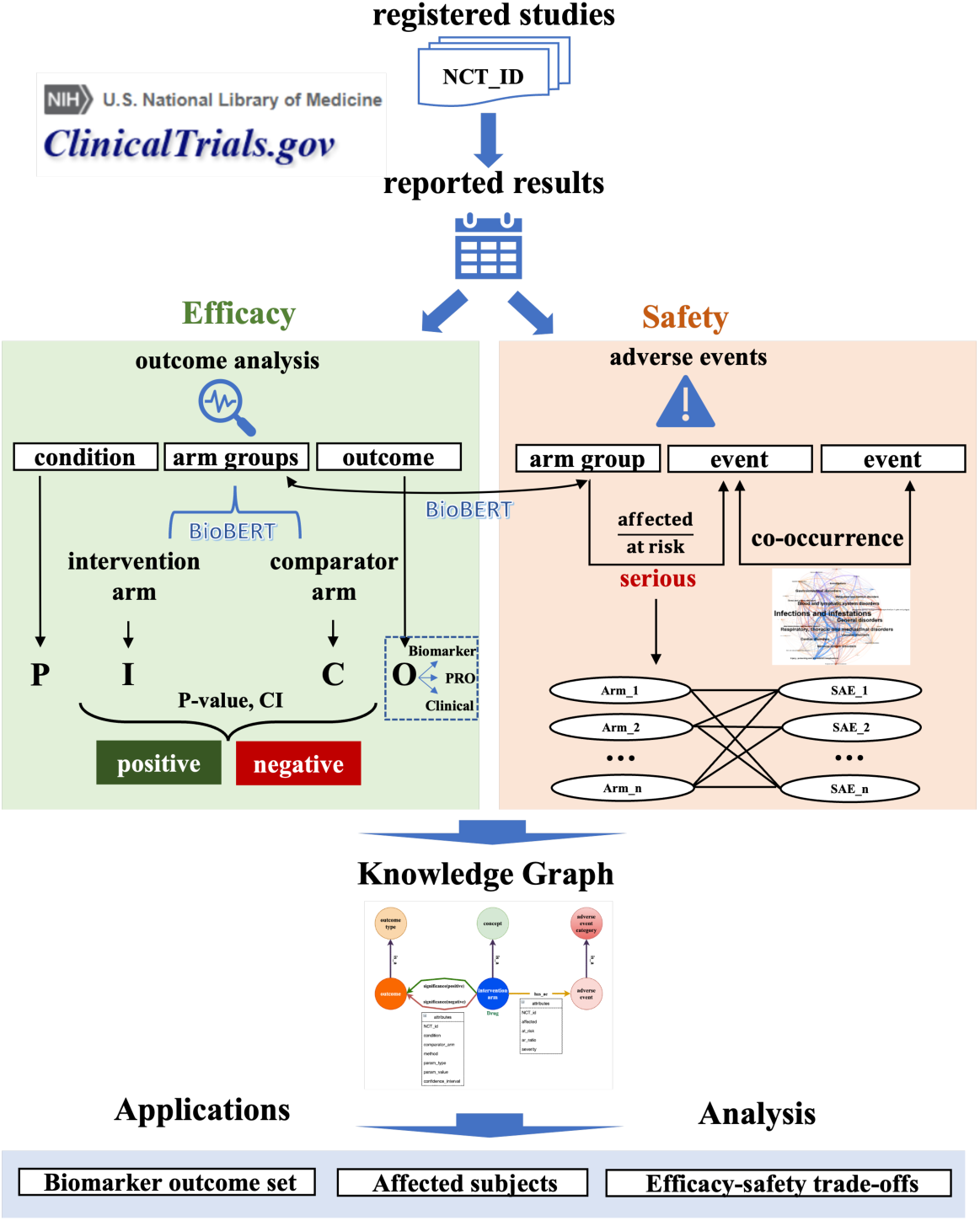
The overall workflow from data acquisition, results processing, to the final knowledge graph construction.

## Methods

### Data source

On December 25^th^ in 2022, We downloaded full registered clinical trials dataset directly from clinicaltrials.gov (https://classic.clinicaltrials.gov/AllPublicXML.zip). The dataset is a compressed zip file containing all the individual XML file of each study named by the NCT id. The total number of XML files of clinical trials downloaded is 437,173. We parsed and transformed each file into a Pandas DataFrame with targeted information. From the reported results section, we focus on efficacy and safety related to the intervention arm group, which are stored under the statistical analysis and adverse events section.

### Arm-level result

In the context of clinical trials, an “arm” refers to a group of subjects receiving a particular intervention, treatment, or control. The term is often used in randomized controlled trials (RCTs), which are considered the gold standard for evaluating the efficacy of different treatments. In a typical RCT, participants are randomly assigned to different arms of the study to minimize the influence of confounding variables and to allow for a fair comparison of the interventions being tested.

### Efficacy

In evidence based medicine, a piece of clinical trial results has to be described as PICO(Population, Intervention, Comparator, Outcome) framework, along with the efficacy(E). In clinical trials, population describes the characteristics of the selection of study subjects, such as age, gender, condition; Intervention represent the treatment or strategy being studied, which can be a drug, a lifestyle or a dietary plan. Comparator represents the strategy for comparison with the intervention, which can a placebo method or a standard care. Outcome describes what is being measure in the study to assess the efficacy and safety of the intervention, which can be symptom relief, biomedical markers, or mortality rate.

The PICO framework is essential for formulating research questions and reviewing literatures for relevant clinical studies. The search for the efficacy of an intervention, it is essential to determine the anticipated corresponding population, comparator, and outcome. In natural language, a clinical trial result can be expressed as: ‘There exists a significant difference between the intervention (I) and the comparator (C) on the outcome (O) in the population (P).

In this study, to evaluate the efficacy of an intervention on an outcome, we searched for the statistical analysis section under each outcome in the study results page. For example, in the study *NCT01050998*, under the outcome measure of the primary outcome ‘*Percentage of Participants Who Achieved Disease Activity Score of 28 Joints Using C-Reactive Protein (DAS28 [CRP]) Response at Day 85 by Region’* exists a list of statistical analysis results. As we can see from an example of results (Fig. 2), the analysis section provides information such as groups, p_value, and confidence interval. With these information we are able to create a relationship between an intervention arm group and the primary outcome.

**Figure 2.**
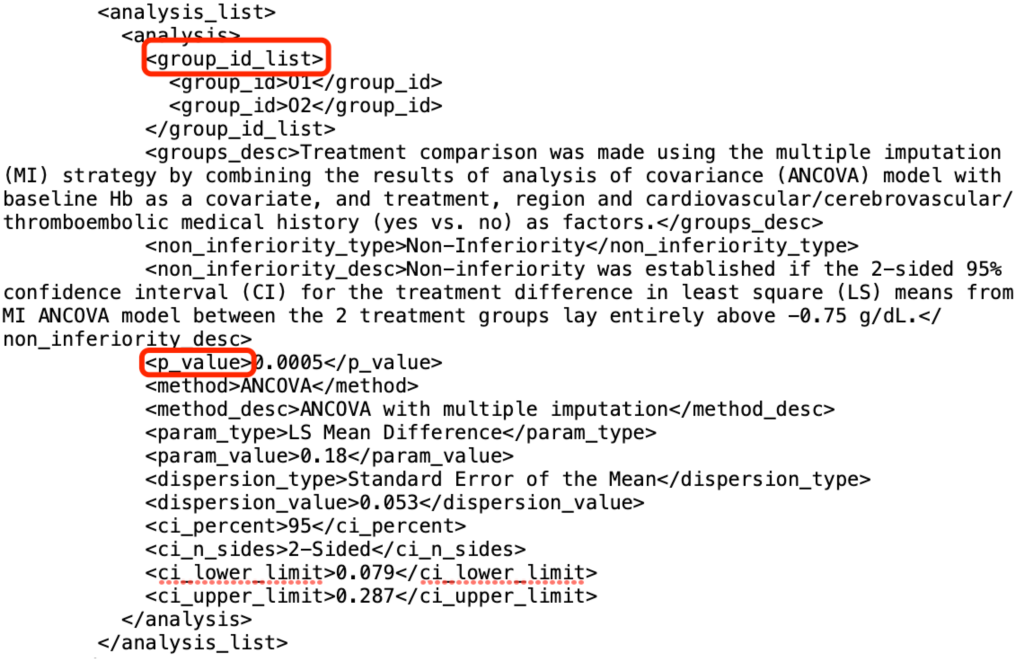
An example of statistical analysis in XML file.

To parse the XML files for efficacy results, we extracted results in the form of relationships between ‘groups’ under each outcome measure and corresponding ‘outcome’: group--outcome. Each group—outcome pair is saved in a row in a Pandas dataframe, along with other columns including other important attributes such as NCT_ID, p-value, statistical method, and confidence interval. An example is shown in Table 1.

**Table 1.** An example of transformed dataframe of efficacy results.

| NCT_ID | groups | outcome_title | p_value |
| --- | --- | --- | --- |
| NCT01049373 | ['Lymphdiaral Basistropfen (HDC)', 'Placebo Solution'] | Change in FFbH-R Between Screening and 2 Weeks | 0.0757 |
| NCT03400800 | ['Inclisiran', 'Placebo'] | Percentage Change in LDL-C From Baseline to Day 510 | <0.0001 |
| NCT02912650 | ['Placebo', 'Ibuprofen 250 mg'] | Pain Intensity Difference on 11-Point Numerical Scale (PID11) | <0.001 |
| NCT02954354 | ['Baloxavir', 'Oseltamivir'] | Percentage of Participants Reporting Normal Temperature at Each Time Point in Adults Randomized to Baloxavir or Oseltamivir | 0.5908 |
| NCT01795547 | ['Aripiprazole', 'Paliperidone'] | Change From Baseline to Week 28 in the 'Interpersonal Relations' QLS Domain Score | 0.07 |

### Statistical significance

To better and directly represent the efficacy of an intervention group on an outcome, we created an automatic rule-based pipeline to summarize analysis attributes for statistical significance and eventually presented as positive vs negative.

(1) check if the result has valid p-value
(2) if no p-value found, check if the result has valid confidence interval
(3) for results with p-value, we label the results as positive if it has a p-value smaller than or equal to 0.05, and negative otherwise
(4) for results without p-value but with confidence interval, we check whether the statistical parameter is a ratio-type parameter (Odds Ratio, Hazard Ratio, etc.) or a difference-type parameter (Mean Difference, Risk Difference, etc.)
(5) for results with a ratio-type statistical parameter, check if the number one (1) is contained between the lower and upper confidence interval limits. If one is not contained, label the result as positive, and negative otherwise
(6) for results with a difference-type statistical parameter, check if the number zero (0) is contained between the lower and upper confidence interval limits. If zero is not contained, label the result as positive, and negative otherwise

The detailed visualized pipeline and corresponding number of results in each step are shown in the result section (Fig. 4).

In the XML clinical trial files downloaded from the website, arm groups are labeled with ‘Experimental’ and ‘Comparator’ in the study design section. But in the statistical analysis section, the intervention arm group is not distinguished from the comparator arm group by using a label. The titles of the groups are also not always consistent with the titles from the study design section, meaning it is not feasible to simply match the labels using the exact strings. Thus, in this part, we utilized a transformer model BioBERT^14^ (‘dmis-lab/biobert-v1.1’) to automatically separate intervention arm group from comparator arm group. BERT, Bidirectional Encoder Representations from Transformers, is a transformers-based deep learning model that was introduced by Google in 2018^15^. It is a language model that is pretrained on a large corpus of texts including Wikipedia. The model allows people to invoke weights (embeddings) of texts from the pretrained model without training on their own. BioBERT is a domain-specific adaptation of the original BERT and pretrained on a large corpus on biomedical texts. With BioBERT, we are able to perform various tasks such as named entity recognition and relation extraction on biomedical problems. In our specific task, we used the version BioBERT-Base v1.1 that is trained on around one million biomedical publications on PubMed.

In this experiment, to prepare the data in the form of the knowledge graph schema, we only kept results with exact two arm groups for comparison between intervention and comparator. Studies without valid labels of ‘experimental’ and ‘comparator’ arm groups from the study design section were excluded.

We first produced semantic embeddings of the arm group titles from both the study design and statistical analysis sections using BioBERT. Then we calculated the similarities of semantic embeddings between the arm groups from different sections. Eventually we labeled the arm group from the statistical analysis section based on the label of the arm group that has the highest similarity score from the study design section. For examples shown in Table 2, the ‘Groups’ column are the original stored groups information from ClinicalTrials.gov. After comparing to columns ‘Intervention’ and ‘Comparator’, new columns ‘Intervention_group’ and ‘Comparator_group’ were created to represent the experimental and comparator groups.

**Table 2.**
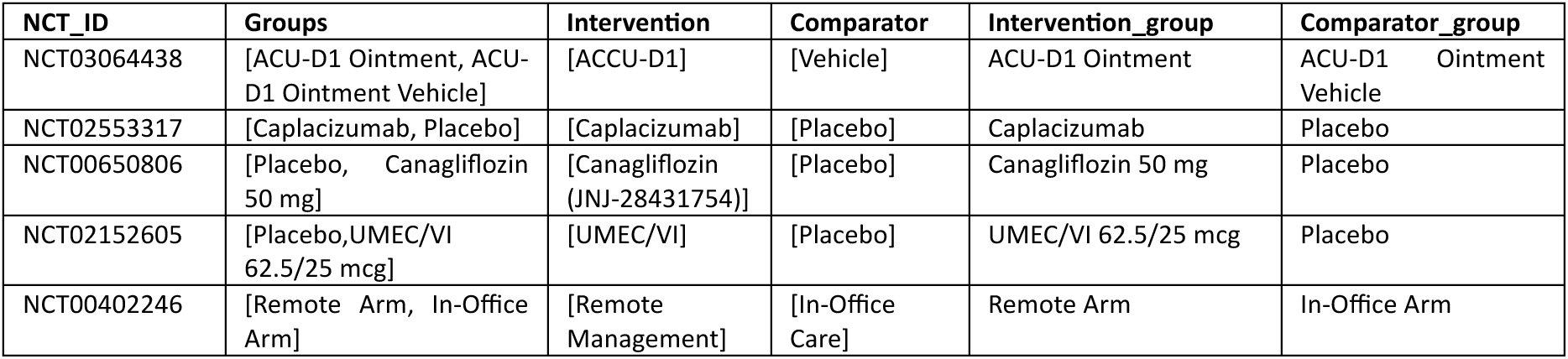
Example results of separation of arm groups into intervention vs. comparator in efficacy results.

### Safety

We integrated reported adverse events from clinicaltrials.gov to represent potential safety harms related to the interventions. In the trial XML file, an adverse event is stored under the <event> section, along with <sub_title> and <count>, recording information including the corresponding group, the number of affected subjects, and the number of subjects at risk. The vast majority of adverse event titles are standardized into the Medical Dictionary for Regulatory Activities (MedDRA^16^) by ClinicalTrials.gov. The parent elements that incorporate the specific events cover health categories such as *‘*Cardiac disorders’, ‘Ear and labyrinth disorders’, etc. The events are also classified as serious events vs. other events, giving users the ability to choose by event severity. In this study, we focus on serious adverse events of which the number of affected subjects is none-zero.

In order to intuitively compare the efficacy and safety, we only kept serious adverse events in the studies that have valid efficacy analysis results (54.8%, 6,429 out of 11,729). On clinicaltrials.gov, not all the expressions of arm groups in adverse events sections are 100% same as the ones in efficacy. In order to integrate efficacy and AE, we first tried using string matching among the arm groups. For arm groups in AE that were not string-matched to an arm group in efficacy, we utilized BioBert again to calculate the semantic similarities and return the closest match. For each adverse event title, we calculated the semantic similarities between the title and all the arm groups in the efficacy section. The closest matched arm group was saved as a unique column in the dataframe, and we used the matched arm group for integrating efficacy and safety. The columns for adverse events we saved in the dataframe include ‘NCT_ID’, ‘arm_title’, ‘event_title’, ‘category’, ‘serious/other’, ‘affected’, ‘at_risk’, and ‘matched group’. Each row can be comprehended as a reported adverse event named ‘event_title’ related to the arm group named ‘matched_group’ along with the number of affected and at-risk participants (Table 3).

- **Events co-occurrence.** In extra, we calculated co-occurrence weights among reported adverse events to provide the which AE is frequently occurred with another AE. We determined the co-occurrence of categories of SAEs by the following considerations: if both two categories have reported ‘affected’ participants in a same trial, and multiplying the ‘affected/at_risk ratio’ from the categories. (∑_(*a*, *b*)∈*s*, *a* ≠b_ *c*(*a*, *b*) × (*r*(*a*) × *r*(*b*))) (c:co-occurrence, r:ratio). This ratio based co-occurrence took the affected/at_risk ratio of SAE into account, representing the probabilistic extent of events co-occurrence.

**Table 3.**
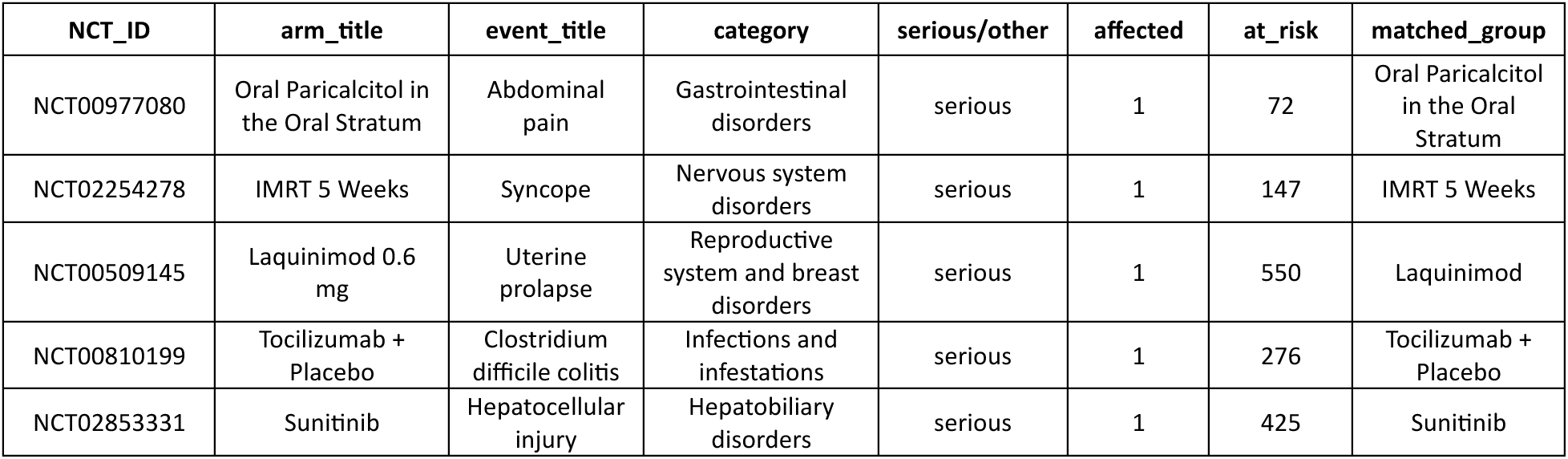
Example adverse events with matched group in efficacy analysis section.

| NCT_ID | arm_title | event_title | category | serious/other | affected | at_risk | matched_group |
| --- | --- | --- | --- | --- | --- | --- | --- |
| NCT00977080 | Oral Paricalcitol in the Oral Stratum | Abdominal pain | Gastrointestinal disorders | serious | 1 | 72 | Oral Paricalcitol in the Oral Stratum |
| NCT02254278 | IMRT 5 Weeks | Syncope | Nervous system disorders | serious | 1 | 147 | IMRT 5 Weeks |
| NCT00509145 | Laquinimod 0.6 mg | Uterine prolapse | Reproductive system and breast disorders | serious | 1 | 550 | Laquinimod |
| NCT00810199 | Tocilizumab + Placebo | Clostridium difficile colitis | Infections and infestations | serious | 1 | 276 | Tocilizumab + Placebo |
| NCT02853331 | Sunitinib | Hepatocellular injury | Hepatobiliary disorders | serious | 1 | 425 | Sunitinib |

### Outcome

#### MeSH extraction

In clinical trial, expressions of outcomes vary across different studies. Thus we processed all the unique outcomes using the MeSH(Medical Subject Headings) extraction tool: NLM Medical Text Indexer(MTI). MTI allows us to extract MeSH terms from natural languages, in our case, outcomes. MeSH represent the simplest representation of biomedical languages for tasks including document retrieval, text classification, etc. For all the MeSH terms extracted, we also recorded the parent terms along with the codings based on the tree structure of MeSH. With MeSH terms of outcomes, we are able to not only standardize outcome titles with different expressions, but also classify outcomes into different clinical categories.

#### Outcome category

In clinical trials, the selection of outcomes has critical impact on the efficacy of the intervention. A study proposed categories of outcomes as Mortality/survival, Physiological/clinical, Life impact, Resource use, and adverse events ^17^. By assumption, with the same intervention, choosing biomarkers as the outcome potentially tend to produce positive results ^18^. Also, biomarkers are usually used as surrogate endpoints that are easier to measure. In contrast, in most cases clinical endpoints are more complicate to measure, and tend to produce negative results. In this study, we classified the outcome titles into three categories: Biomarker, Patient-reported outcome (PRO), and clinical endpoint.

- **Biomarker.** We took the advantage of MeSH to identiy biomarker outcomes. We first manually went through the MeSH tree to discover possible biomarker-related terms. We found that the majority of the terms we discovered (such as ‘E01.370.376.537.250-Brain Cortical Thickness’, ‘D12.776-Proteins’, and ‘D10.251-Fatty Acids’) belong to the root category D(Chemicals and Drugs) and E(Analytical, Diagnostic and Therapeutic Techniques, and Equipments). Thus, we preliminarily selected the root categories D and E as the biomarker identifier in outcomes. If any of the MeSH terms extracted from an outcome title starts with D or E, we labeled the outcome as biomarker. We excluded ‘E05.318.308.980-Surveys and Questionnaires’ as it is a PRO related MeSH term. 40,525 results with unique 14,217 biomarker outcomes were identified.
- **PRO.** Patient-reported outcomes(PROs) are used to report the status of a patient’s health condition from the patient’s persepective^19^. Research identified PROs and inspected the inclusion of PROs in registered clinical trials from 2007 to 2013 and found 27% of the trials used one or more PRO measures^20^. Recently, the usage of PROs in novel artificial intelligence(AI) clinical trials was assessed by researchers (152/627 trials)^19^. Both previously-mentioned studies used PRO databases such as PROQOLID and GEM to match PROs in registered clinical trials. One of the major limits of exact text-matching for outcomes in registered clinical trials is that the writings or expressions are natural languages, meaning they can be inconsistent. In this study, we simplified the searching words and took advantage of the extracted MeSH terms. To identify PRO outcomes, we applied keyword matching on columns ‘outcome_title’ and ‘outcome_mesh’. The keywords are ‘Survey’, ‘Questionnaire’, ‘Patient Reported’, ‘Patient-Reported’, ‘Scale’, and ‘Score’. We labeled the outcome as PRO if any lowercase of the keywords was matched in the outcome string. We eventually found 33,547 results with 9,820 unique PRO outcomes.
- **Clinical endpoint.** In this study, we defined the disease-related outcomes as the clinical endpoint outcomes. The root category C from MeSH tree represents ‘Diseases’. We also added F03 (Mental Disorders) as a target disease-related category. If any of the MeSH terms extracted from an outcome title starts with ‘C’ and ‘F03’, we labeled the outcome as clinical endpoint. 23,557 results with 7,837 unique clinical endpoint outcomes were identified.

In short summary, 25,758 (61.03%) unique outcomes were successfully identified as biomarker, PRO, or clinical endpoint, contained in 76,575 (63.83%) efficacy results. We noticed that there existed differences of the coverage of outcome categories between primary outcomes and secondary outcomes. Table 4 shows the detailed coverage numbers of biomarkers, PROs, and clinical endpoint outcomes.

**Table 4.** Coverage statistics of Identified categories in Primary and Secondary outcomes (‘result’ means numbers in efficacy results, ‘outcome’ means numbers of unique outcomes in efficacy results)

|  | Primary (result) | Secondary (result) | Primary (outcome) | Secondary (outcome) |
| --- | --- | --- | --- | --- |
| **Total** | 23,252 | 92,966 | 10,624 | 30,408 |
| Biomarker | 9,321 (40.09%) | 29,931 (32.20%) | 4,105 (38.64%) | 9,702 (31.91%) |
| PRO | 4,378 (18.83%) | 27,838 (29.94%) | 2,072 (19.50%) | 7,501 (24.67%) |
| Clinical | 4,133 (17.77%) | 18,700 (20.11%) | 2,080 (19.58%) | 5,537 (18.21%) |
| **coverage** | 14,388 (61.88%) | 59,738 (64.26%) | 6,609 (62.21%) | 18,429 (60.61%) |

### Knowledge graph construction

A knowledge graph (KG) is a specialized graph-based data structure used for representing a collection of knowledge, entities, and their relationships. In this study, we stored the clinical trial entities (outcome, arm group, adverse event, etc.) and their relationships in a knowledge graph for visual representation and complex data retrieval queries.

We created 3 main node types: intervention_arm, outcome, and adverse_event:

- **Intervention_arm**. The intervention_arm node is interventional arm group from the statistical analysis result section. The relationship representing the efficacy of results is either ‘positive’ or ‘negative’, with intervention_arm as starting node and outcome as ending node, built based on the columns ‘intervention_group’, ‘outcome_title’, and ‘siginificance’ from each efficacy result row. ‘Positive’ relationships are shown in green and ‘negative’ relationships are shown in red. Some intervention_arm nodes are connected to a concept node that saves the mapped MeSH term of the intervention_arm title.
- **Outcome**. The outcome node is clinical outcome used in each study. The outcome node can be connected to an outcome_type node to exhibit the types of an outcome, including biomarker, PRO, and clinical endpoint.
- **Adverse_event**. The adverse_event node is reported serious adverse event in each clinical trial. For adverse events, we built the relationship ‘has_ae’ from the column ‘intervention_arm’ to ‘event_title’ from each adverse event result row. Each adverse_event node is connected to an adverse_event_category node, representing the parent biomedical categories of the adverse event name.

We loaded the created nodes and relationships along with their attributes into the graph database Neo4j (database version==5.3.0).

To summarize, we presented a pipeline to construct a knowledge graph to represent arm-level clinical trial efficacy and safety results. Compared to existing large scale trial databases such as AACT and CTKG, we curated a result database containing identified comparable arm groups in results section. We also integrated efficacy and safety results by connecting corresponding arm groups, providing an evidence dataset for evaluating clinical interventions. We constructed a knowledge graph based on the dataset, offering a data infrastructure for further analysis and applications. However, current limitations include: (1) In efficacy results, multiple testings are not adjusted yet. (2) Multiple arms were not included and classified as interventional and comparator. (3) Current dataset only covers clinical trials with both efficacy and safety results.

## Data Records

### Knowledge graph schema

The knowledge graph schema exhibits the most important nodes, relationships, and attributes when evaluating clinical trial results. Compared to the sole ‘has’ relationship in the CTKG, in order to better represent the RESULTS across different studies, we saved all the study-related information of the results in different relationships instead of unique nodes, based on the efficacy and adverse event (Fig. 3). For efficacy, the schema shows the significance, the comparator arm, the parameter value, NCT_id, and condition. For safety, the schema shows the affected/at_risk ratio, NCT_id, and severity. Also, for all the 3 main node types, we applied different techniques described in the method section to give each node type a standardized or taxonomic categories for data integrity and normalization. Table 5 shows the counts of node types and relationships.

**Figure 3.**
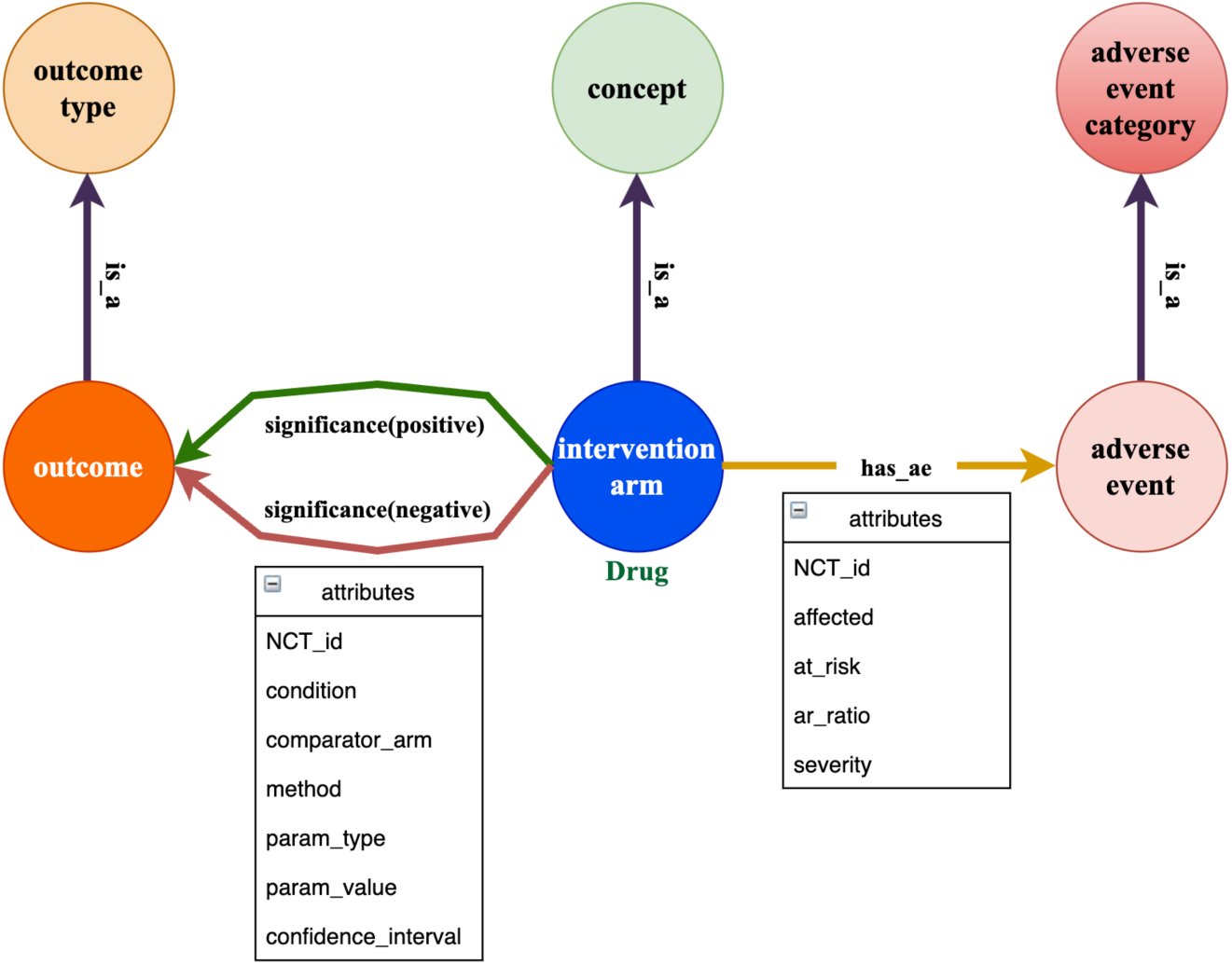
An overview of the schema of arm-level efficacy and safety knowledge graph.

**Figure 4.**
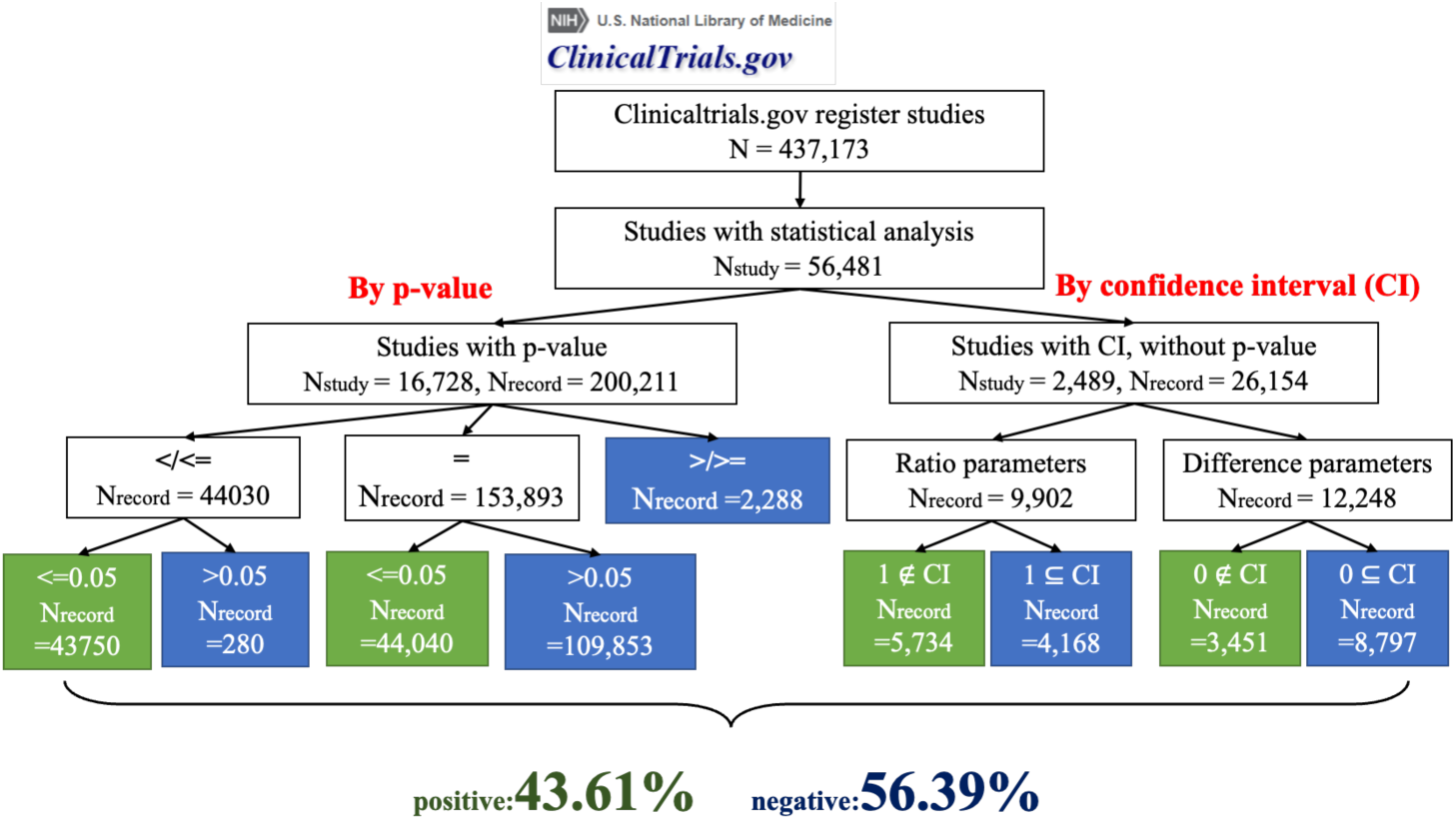
The pipeline and numbers of classification of positive vs. negative results.

**Table 5.** Statistics nodes and efficacy and safety relationships. The relation count of efficacy is the number of results with valid intervention group, comparator group and statistical significance. The relation count of safety is the number of serious adverse events with valid arm group and event title.

| Node type 0 | Count 0 | Node type 1 | Count 1 | Relation name | Relation count |
| --- | --- | --- | --- | --- | --- |
| Intervention_arm | 10,364 | Outcome | 41,708 | positive/negative | 119,968 |
| Intervention_arm | 10,364 | Adverse_event | 40,408 | has_ae | 803,052 |
| Intervention_arm | 10,364 | Concept | 1439 | is_a | 49,764 |
| Outcome | 41,708 | Outcome type | 3 | is_a | 76,575 |
| Adverse event | 40,408 | Adverse event category | 28 | is_a | 803,052 |

### Efficacy

To determine the efficacy or statistical significance of an intervention on an outcome, we used p-values and confidence intervals. However, these fields lacked clarity and uniformed formatting. To transform the uncleaned string data into a computable format for our rule-based algorithm, we removed blanks in the strings, transformed the numbers to numeric, and processed the mathematical operators in standard representations. After using the classification pipeline, we introduced in the method section, we eventually distinguished 56.39% statistically negative results, and 43.61% statistically positive results.

### Safety

We investigated the number of affected subjects of SAEs related to the arm groups. In this dataset, a total of 2,106,063 subjects (not necessarily unique individual subjects) were affected by SAEs, and 3,538,554 were at risk. Table 6 shows the top 10 intervention mesh terms of arm groups that are associated with the greatest number of affected subjects. Table 6 only included SAEs with valid one-to-one mesh terms and types. Note that this is not a causal relationship between SAE and arm group. The reasons of appearances of SAEs can be related to the subjects’ personal health condition, trial design, disease complications, etc. Table 7 lists the top SAE categories that are associated with the largest numbers of affected subjects.

**Table 6.**
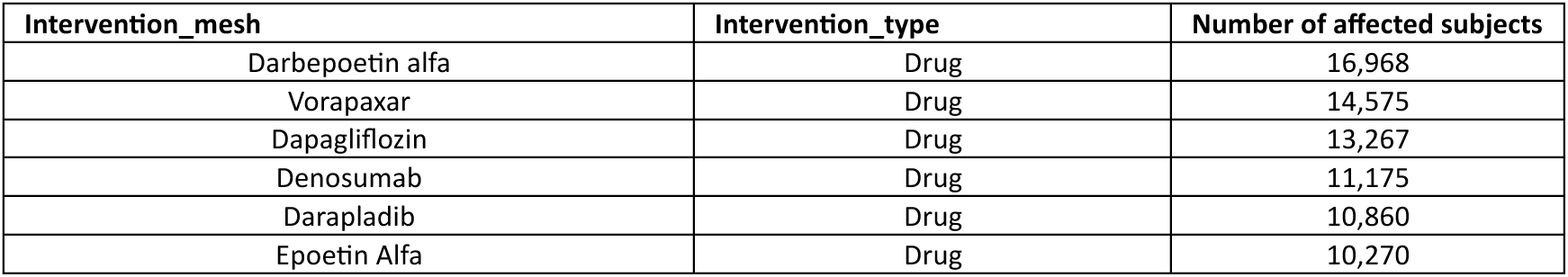
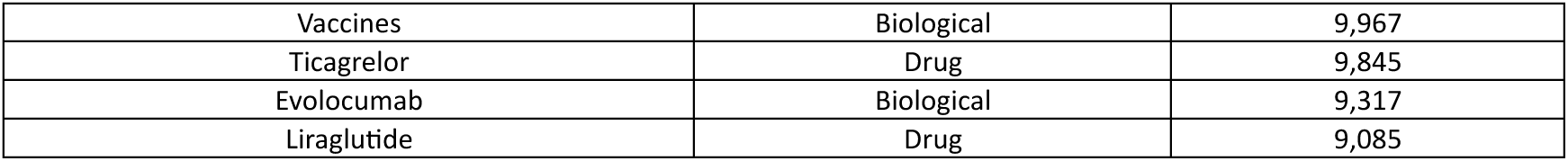
Numbers of affected subjects by different arm groups.

**Table 7.** Numbers of affected subjects by SAE categories.

| category name | Number of affected subjects |
| --- | --- |
| Infections and infestations | 294,602 |
| Cardiac disorders | 286,035 |
| General disorders | 195,018 |
| Gastrointestinal disorders | 168,440 |
| Respiratory, thoracic and mediastinal disorders | 147,184 |
| Neoplasms benign, malignant and unspecified (incl cysts and polyps) | 132,976 |
| Nervous system disorders | 129,127 |
| Injury, poisoning and procedural complications | 104,784 |
| Vascular disorders | 93,766 |
| Blood and lymphatic system disorders | 78,164 |

Also note that this is only the safety results that from studies having efficacy results at the same time, for the purpose of tracking original study for integrating efficacy and safety analysis. We imported the co-occurrence matrix to a network visualization software Gephi^21^ and output the co-occurrence map of SAE categories (Fig. 5). In the co-occurrence map, each node represents an adverse event category. Each edge represents the existence of co-occurrence between two categories. The width of edges represents the weighted co-occurrence scores. The size of nodes represents the sum of the scores. Table 8 lists the top co-occurrence edges between SAE categories, with corresponding calculated weights. The table compares the degrees of co-occurrence of SAE categories. With the co-occurrence algorithm (AR ratio), the SAE category ‘Infections and infestations’ has the highest degree of co-occurrence, meaning it is the most frequently and strongly accompanied by other SAEs.

**Figure 5.**
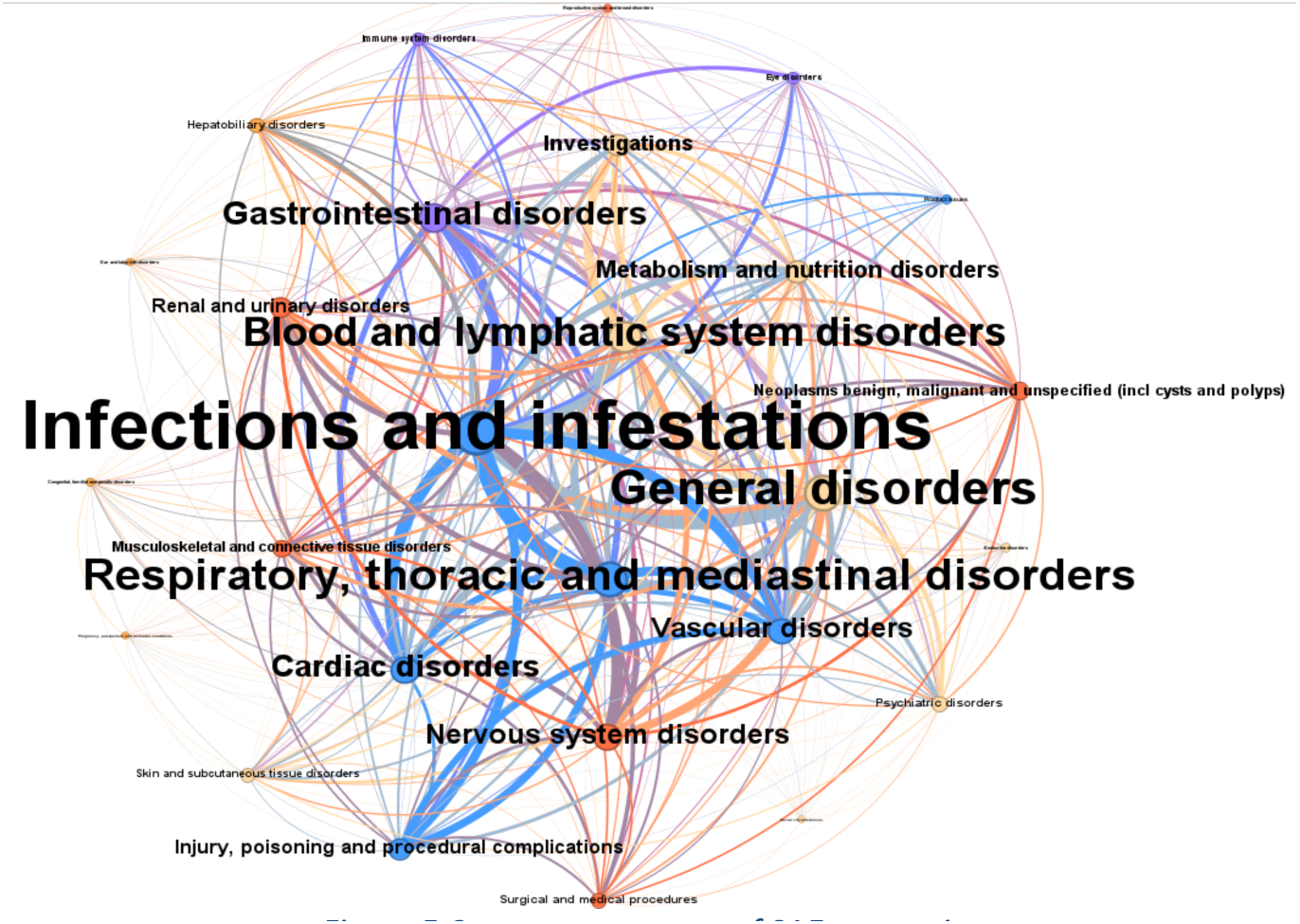
Co-occurrence map of SAE categories.

**Table 8.**
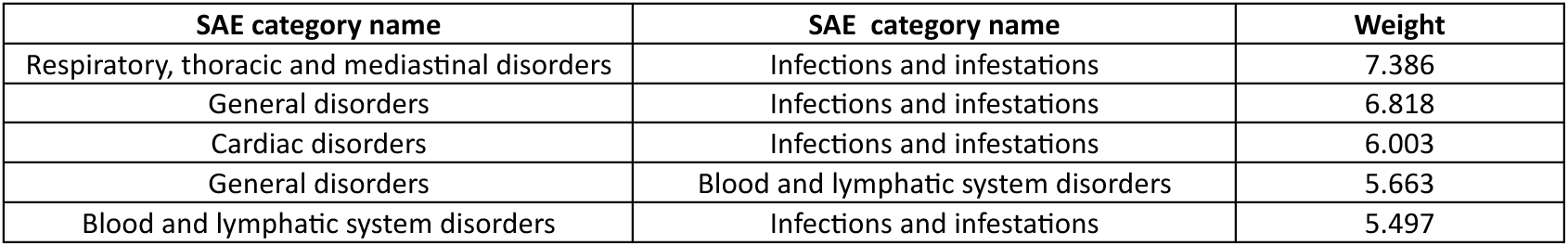
Top co-occurrence edges with ratio weights.

| SAE category name | SAE category name | Weight |
| --- | --- | --- |
| Respiratory, thoracic and mediastinal disorders | Infections and infestations | 7.386 |
| General disorders | Infections and infestations | 6.818 |
| Cardiac disorders | Infections and infestations | 6.003 |
| General disorders | Blood and lymphatic system disorders | 5.663 |
| Blood and lymphatic system disorders | Infections and infestations | 5.497 |

### Knowledge graph: Antibiotic and infection

In order to present the functionality of evaluating efficacy and safety of an intervention based on the constructed knowledge graph, we were consulted by Na He, a pharmacist from the Department of Pharmacy, Peking University Third Hospital. Na was interested in adverse events related to infections while antibiotic ‘imipenem/cilastatin’ being used to treat infections. We queried the knowledge graph, retrieved efficacy results by matching the ‘intervention_group’ and ‘intervention_MeSH’ with ‘imipenem’ or ‘cilastatin’, and retrieved adverse event results by matching the group and adverse events title ‘event_title’ and parent category ‘category’ with the keyword ‘infection’. The trade-offs between efficacy and safety are crucial in clinical studies, especially the population condition and potential adverse events belonging to the same clinical condition. The queried relationships are shown in Fig. 6 and detailed SAE results are shown in Table 9. The infection-related SAEs give us evidence which types of infections should be paid attention when treated with imipenem/cilastatin.

**Figure 6.**
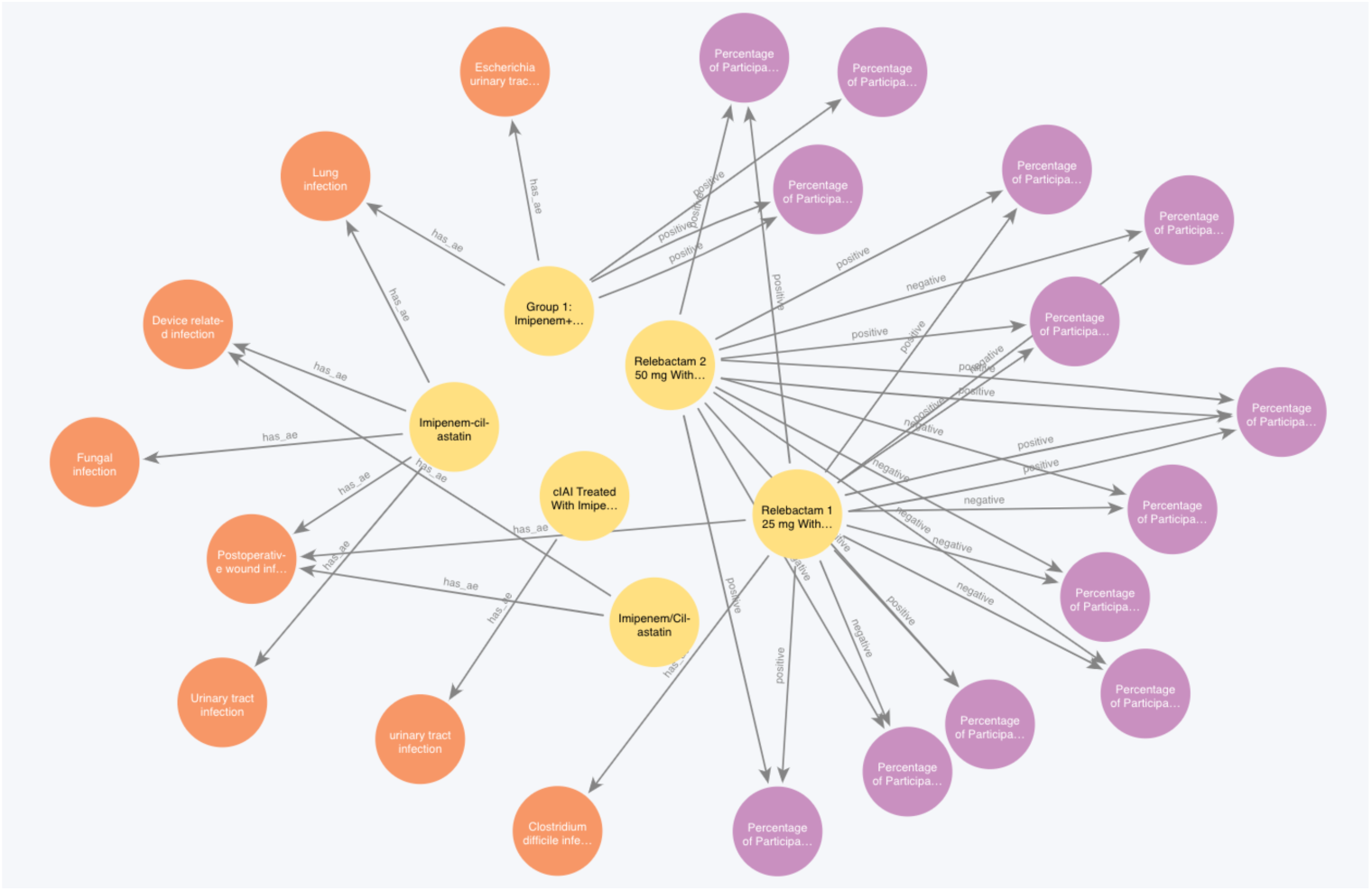
An example KG that shows the case of infection-related adverse events while intervening with the antibiotic imipenem/cilastatin to treat infections. Orange nodes are SAEs, yellow nodes are interventional arm groups, purple nodes are outcomes, visualized in Neo4j Bloom.

**Table 9.**
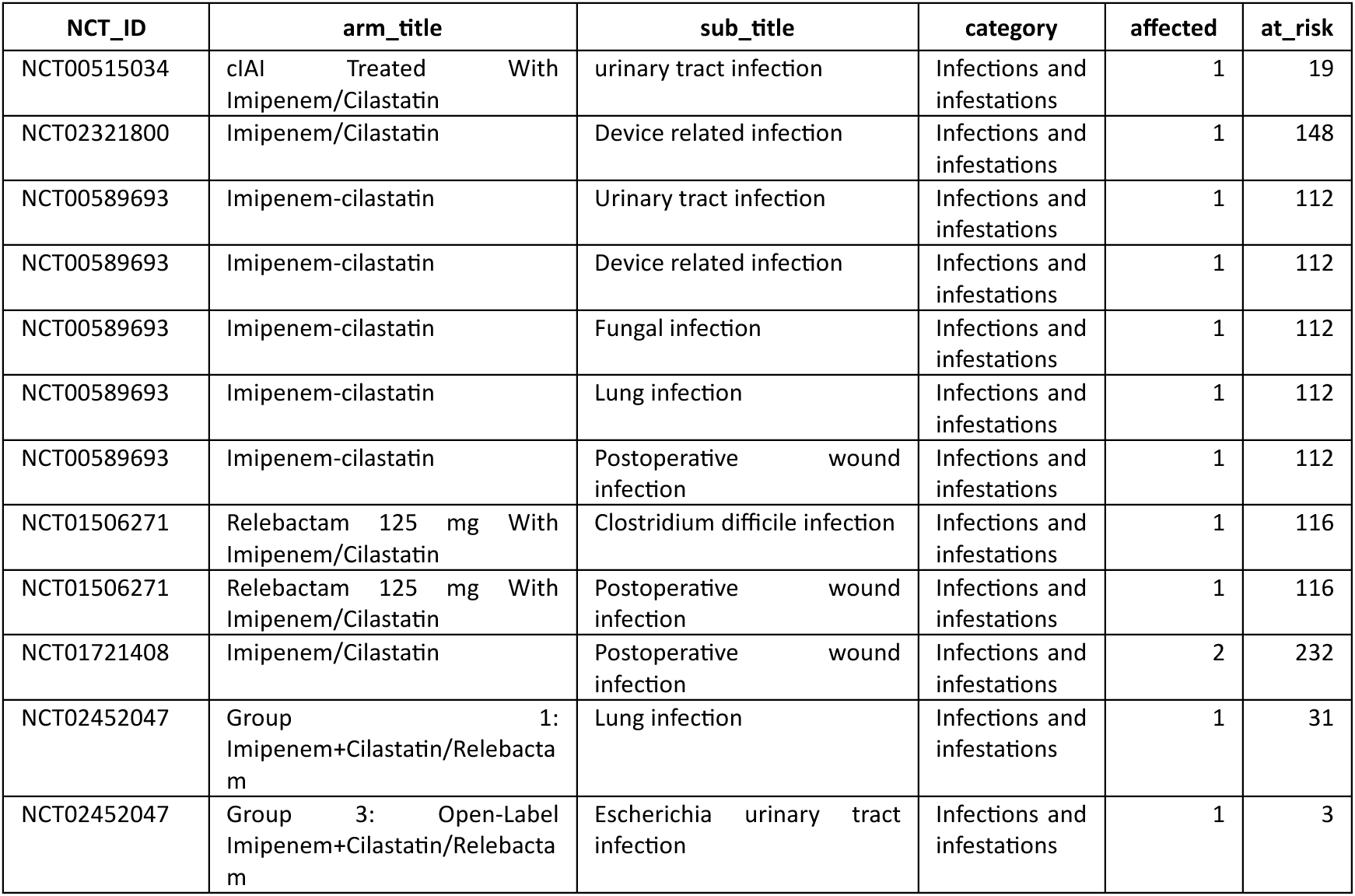
Infection-related serious adverse events with antibiotic ‘imipenem/cilastatin’.

| NCT_ID | arm_title | sub_title | category | affected | at_risk |
| --- | --- | --- | --- | --- | --- |
| NCT00515034 | cIAI Treated With Imipenem/Cilastatin | urinary tract infection | Infections and infestations | 1 | 19 |
| NCT02321800 | Imipenem/Cilastatin | Device related infection | Infections and infestations | 1 | 148 |
| NCT00589693 | Imipenem-cilastatin | Urinary tract infection | Infections and infestations | 1 | 112 |
| NCT00589693 | Imipenem-cilastatin | Device related infection | Infections and infestations | 1 | 112 |
| NCT00589693 | Imipenem-cilastatin | Fungal infection | Infections and infestations | 1 | 112 |
| NCT00589693 | Imipenem-cilastatin | Lung infection | Infections and infestations | 1 | 112 |
| NCT00589693 | Imipenem-cilastatin | Postoperative wound infection | Infections and infestations | 1 | 112 |
| NCT01506271 | Relebactam 125 mg With Imipenem/Cilastatin | Clostridium difficile infection | Infections and infestations | 1 | 116 |
| NCT01506271 | Relebactam 125 mg With Imipenem/Cilastatin | Postoperative wound infection | Infections and infestations | 1 | 116 |
| NCT01721408 | Imipenem/Cilastatin | Postoperative wound infection | Infections and infestations | 2 | 232 |
| NCT02452047 | Group 1: Imipenem+Cilastatin/Relebactam | Lung infection | Infections and infestations | 1 | 31 |
| NCT02452047 | Group 3: Open-Label Imipenem+Cilastatin/Relebactam | Escherichia urinary tract infection | Infections and infestations | 1 | 3 |

### Technical Validation

We introduced the full pipeline from data collection, data processing to the final knowledge graph in details. The knowledge graph and original dataset is transparent in the following aspects: 1) The origin of data comes from ClinicalTrials.gov, which is an open-access platform that everyone is able to download the full structured dataset. 2) The process of formation of the knowledge graph is reproducible. The codes will be available to access publicly. 3) Users are able to adjust the knowledge graph based on provided codes and sub-datasets, providing the flexibility to alter the data based on their own research interests. Next, we present three case studies to validate the **usability** for exploratory analysis, **consistency** with external resources, and **feasibility** for real scientific problems.

### Case study 1: Positive vs. negative efficacy results across types of conditions, interventions, and outcomes

We investigated the ratio of positive and negative efficacy results across different dimensions (Table 10). For technical validation, we also provide a biomarker outcome list for each condition. Each list contains a list of biomarker-type outcomes with corresponding number of positive/negative efficacy results. This dataset provides clinical researchers and practitioners surrogate endpoints for measurement when it is infeasible, unethical, or ineffective to directly evaluate clinical endpoints. This case confirmed the usability of the dataset and users are capable of conduct exploratory data analysis from every dimension.

**Table 10.** Distribution of positive and negative efficacy results across different dimensions.

|  | positive | negative | positive_rate | negative_rate |
| --- | --- | --- | --- | --- |
| **<u>Outcome type</u>** |  |  |  |  |
| Biomarker | 14166 | 15148 | 48.33% | 51.67% |
| Patient-reported outcome | 12647 | 20900 | 37.70% | 62.30% |
| Clinical endpoint | 9820 | 13757 | 41.65% | 58.35% |
| **<u>Intervention type</u>** |  |  |  |  |
| Drug | 43521 | 54294 | 44.49% | 55.51% |
| Biological | 5507 | 7168 | 43.45% | 56.55% |
| Radiation | 57 | 143 | 28.50% | 71.50% |
| Behavioral | 1019 | 2375 | 30.02% | 69.98% |
| Genetic | 18 | 53 | 25.35% | 74.65% |
| Other | 511 | 966 | 34.60% | 65.40% |
| Device | 1223 | 1919 | 38.92% | 61.08% |
| Dietary Supplement | 155 | 389 | 28.49% | 71.51% |
| Procedure | 147 | 380 | 27.89% | 72.11% |
| Combination Product | 71 | 48 | 59.66% | 40.34% |
| Diagnostic Test | 1 | 3 | 25.00% | 75.00% |
| **<u>Health Problems</u>** |  |  |  |  |
| Infections | 2640 | 3660 | 41.90% | 58.10% |
| Neoplasms | 1152 | 1929 | 37.39% | 62.61% |
| Musculoskeletal Diseases | 4338 | 5108 | 45.92% | 54.08% |
| Digestive System Diseases | 1007 | 1352 | 42.69% | 57.31% |
| Stomatognathic Diseases | 544 | 660 | 45.18% | 54.82% |
| Respiratory Tract Diseases | 4520 | 6337 | 41.63% | 58.37% |
| Otorhinolaryngologic Diseases | 520 | 644 | 44.67% | 55.33% |
| Nervous System Diseases | 2090 | 5086 | 29.12% | 70.88% |
| Eye Diseases | 478 | 940 | 33.71% | 66.29% |
| Urogenital Diseases | 1438 | 1461 | 49.60% | 50.40% |
| Female Urogenital Diseases and Pregnancy Complications | 1614 | 1621 | 49.89% | 50.11% |
| Cardiovascular Diseases | 2071 | 2647 | 43.90% | 56.10% |
| Hemic and Lymphatic Diseases | 434 | 639 | 40.45% | 59.55% |
| Congenital, Hereditary, and Neonatal Diseases and Abnormalities | 1398 | 1325 | 51.34% | 48.66% |
| Skin and Connective Tissue Diseases | 3539 | 3870 | 47.77% | 52.23% |
| Nutritional and Metabolic Diseases | 6091 | 5331 | 53.33% | 46.67% |
| Endocrine System Diseases | 3546 | 3190 | 52.64% | 47.36% |
| Immune System Diseases | 4425 | 6122 | 41.96% | 58.04% |
| Pathological Conditions, Signs and Symptoms | 5483 | 7817 | 41.23% | 58.77% |
| Occupational Diseases | 8 | 174 | 4.40% | 95.60% |
| Chemically-Induced Disorders | 107 | 150 | 41.63% | 58.37% |
| Wounds and Injuries | 131 | 331 | 28.35% | 71.65% |
| Mental Disorders | 2408 | 4575 | 34.48% | 65.52% |

### Biomarker outcome set

We provide a dataset of biomarker outcomes with the most positive and negative efficacy results under each health problem. The dataset is provided in Supplementary Table 1. Note that the outcomes are not standardized by concept, meaning there might be variations of same outcomes that are not summed together. Besides, this example shows the top biomarker outcomes associated with health problems. In the full dataset, outcomes should be associated with the original condition name.

### Case study 2: Top SAE categories comparison with FDA data

US Food and Drug Administration (FDA) manages an adverse event reporting database (FDA’s Adverse Event Reporting System, FAERS) intending to support post-marketing safety surveillance for drug and biomedical products. Compared to RCT results on CT.gov, FAERS receive reports directly from healthcare professionals and consumers.

Simon Lafosse visualized the distribution of adverse events by body systems from FAERS^22^. The visualization shows that systems such as ‘General Disorders’, ‘Injury, Poisoning’, and ‘Skin’ have most of the reported AE cases from April 2018 to March 2019. Inspired of this work, with results data of ClinicalTrials.gov in hand, we inspected the sum of numbers of affected subjects under each SAE category between 2018 and 2019 (Fig. 7). In this case, we included all the possible interventions (top 20 in Simon Lafosse’s), and also visualized the distribution of SAE by categories. ‘Infections and infestations’ and ‘Cardiac disorders’ are related to 38,929 and 38,000 reported affect subjects in this dataset. This case validates that our dataset can be used to analyze SAE in clinical trials on a large scale. Also compared to the analysis of FDA data, we found some common top condition categories such as ‘Nervous system disorders’, and ‘Infections and infestations’. There is great potential of deeper analysis of SAE based on this dataset since it has multiple levels of meta-data including co-occurrence weights, mapped categories, corresponding efficacy results, etc. This case shows the consistency of ability to analyze adverse events by condition category from ClinicalTrials.gov data, similar to other analysis based on FDA data.

**Figure 7.**
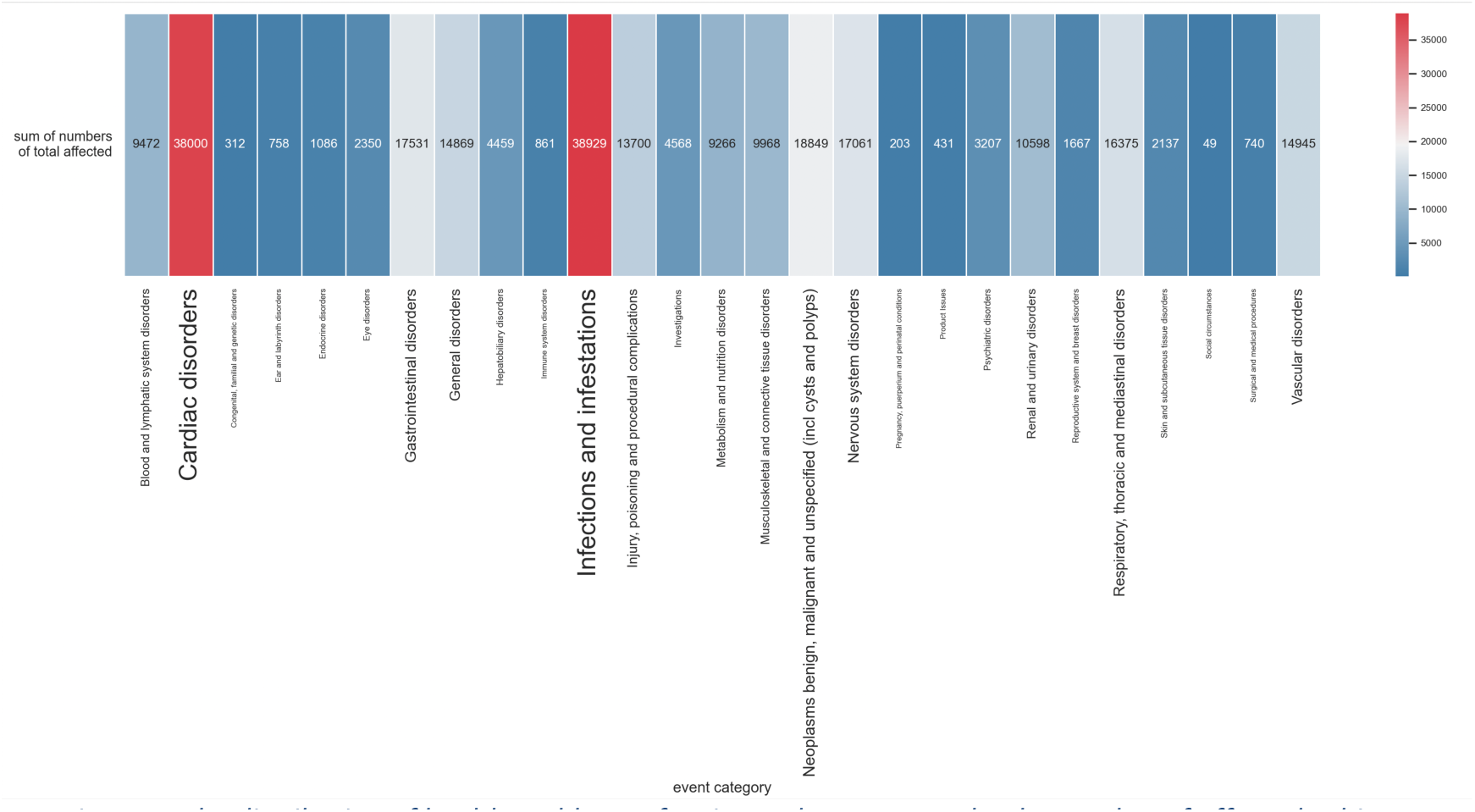
The distribution of health problems of serious adverse events by the number of affected subjects.

### Case study 3: Interventions with same condition and SAE category

Multimorbidity and comorbidity are vital challenges to medical practice^23,24^. We attempt to find interventional arm groups found having associated SAE categories while used to treat the same category. In this case, we selected ‘Infections and Infestations’ as the category since it is the category that is associated most trial subject. Fig. 8 illustrates the KG retrieval of above circumstance, with yellow nodes are intervention arm groups, orange nodes are SAEs, and purple nodes are outcomes. This case validates that with real medical practice problem, this knowledge graph has much potential to provide reliable, large-scale, and human-readable evidence.

**Figure 8.**
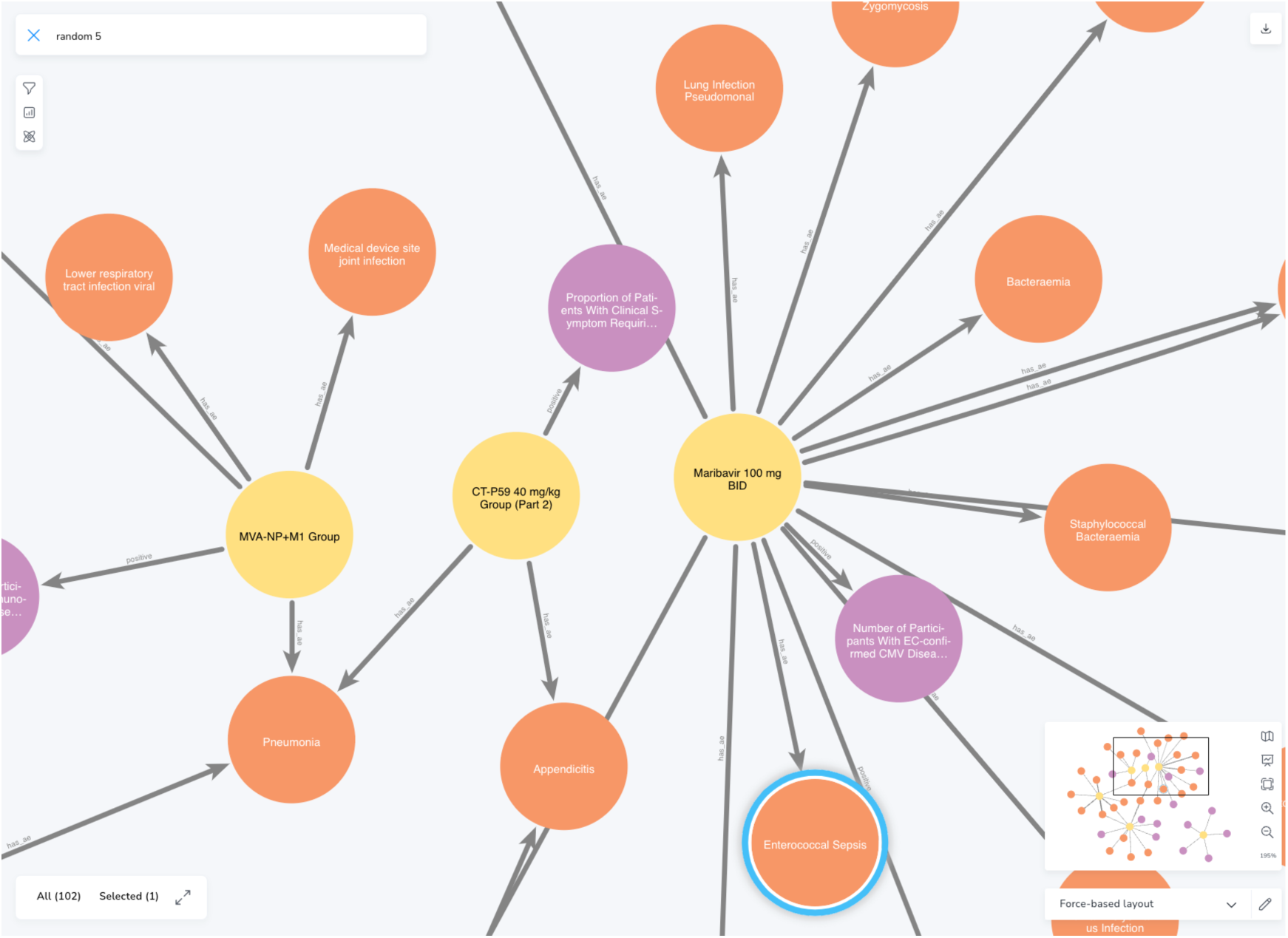
KG example in which both efficacy and safety outcomes are infections, visualized in Neo4j Bloom.

We retrieved the arm groups and corresponding mesh terms and saved them to provide an intervention set for researchers who want to find interventions that are tested positive for efficacy and are observed with SAEs from the same condition category at the same time. This knowledge graph can also be used to retrieve for other conditions. The full arm groups set and corresponding mesh terms are in the following:

### Arm groups set

*[’Vaniprevir 24 Week Arm’, ‘Rabipur Group’, ‘7vPnC After Infant Series Dose 3’, ‘MK-3415 + SOC’, ‘Nevirapine QD+BID’, ‘Vaccine’, ‘V710 60 µg’, ‘CD24Fc’, ‘Enhanced Treatment’, ‘Priorix-Tetra Group’, ‘CCI Vaccine’, ‘Group D: TDV Lyophilized’, ‘Control’, ‘HIV+/Cervarix Group’, ‘rhNGF 20 µg/ml’, ‘Telaprevir 24 Week+Peg-IFN-alfa-2a,RBV 48 Week’, ‘V419’, ‘Placebo + INFANRIX Hexa’, ‘QIVc (≥4 to <18 Years)’, ‘Baricitinib + SOC’, ‘ADV-TDF’, ‘Ziprasidone’, ‘Reltecimod (AB103) 0.5 mg/kg’, ‘Cohort 1-Engerix-B/Prevnar-Hiberix Group’, ‘Arm 1: Metronidazole Plus Miconazole’, ‘Neutrolin Arm’, ‘SB213503 Lot 1 + M-M-R Group’, ‘Azithromycin’, ‘DTG + RPV’, ‘Twinrix Group’, ‘IVV Vaccine’, ‘QIVc (≥18 Years)’, ‘7vPnC After the Infant Series’, ‘ABC/ DTG/ 3TC’, ‘Immediate Introduction of Everolimus’, ‘Baloxavir Marboxil’, ‘PTX+ART’, ‘RotaTeq™’, ‘4200mg Bamlanivimab’, ‘MMR Group’, ‘DTaP (Catch-up 7vPnC)’, ‘CAB LA+RPV LA (Q4W)’, ‘Placebo– Asia’, ‘Immediate ART’, ‘Telaprevir 12 Week+Peg-IFN-alfa-2a,RBV 48 Week’, ‘Letermovir’, ‘Cyclosporine’, ‘Bimekizumab (PPS)’, ‘Maribavir 400 mg’, ‘V501’, ‘Investigational Device’, ‘Oral Care With Influenza and Pneumococcal Vaccines’, ‘MK-6072 + SOC’, ‘Placebo for 24 Weeks’, ‘TMC435 150mg 12Wks PR24/48’, ‘Cohort 2’, ‘Td Adsorbed Vaccine’, ‘Baloxavir’, ‘Pneumosil’, ‘CT-P59 40 mg/kg Group (Part 2)’, ‘Test’, ‘Delafloxacin’, ‘Zoster-022 GSK1437173A 70-79YOA Group’, ‘Group 1: QIV-HD’, ‘Phase 3: 700 mg Bamlanivimab + 1400 mg Etesevimab’, ‘Cohort 3’, ‘Azithromycin + Chloroquine’, ‘Maribavir 100 mg BID’, ‘Camostat Mesilate’, ‘7vPnC After 2-Dose Infant Series’, ‘V260 With Staggered or Concomitant EPI’, ‘Anaferon for Children’, ‘M72AS01 Group’, ‘Double-Blind Period– Vicriviroc 20 mg Plus an ART Regimen’, ‘MEDI8897 50 mg’, ‘MK-8237’, ‘TIV (Elderly)’, ‘Peginterferon Alfa-2a + Ribavirin’, ‘Clinically Driven Monitoring (CDM)’, ‘Telaprevir 12 Week +Peg-IFN-alfa-2a,RBV 24 Week’, ‘Raltegravir 800 mg q.d.’, ‘V419 Lots A, B, and C Combined’, ‘MK-3415A + SOC’, ‘Combined Patients Receiving 74 Gy RT’, ‘SYN-004’, ‘Nevirapine’, ‘Cohort 3 (Placebo, 2-5 Yrs)’, ‘Camostat Mesylate’, ‘Telaprevir 8 Week, PBO 4 Week+Peg-IFN-alfa-2a, RBV 24/48 Week’, ‘DRV/COBI+ FTC/TDF (Control) (Baseline to Switch)’, ‘TDV Lot 3’, ‘qHPV Vaccine in Base Study’, ‘Telaprevir 12 Week+Peg-IFN-alfa-2a,RBV 24 Week’, ‘Double-Blind Period– Vicriviroc 30 mg Plus an ART Regimen’, ‘MVA-NP+M1 Group’, ‘Maternal Chloroquine Prophylaxis’, ‘FluLaval™ Quadrivalent Group’, ‘4_rhNGF10_Phase 2_treatment’, ‘V114’, ‘High-Dose Quadrivalent Influenza Vaccine (QIV-HD)’, ‘GSK1437173A Group’, ‘PR5I’, ‘Stopped Cotrimoxazole Prophylaxis’, ‘GSK1437173A Overall Ages Group’, ‘AZD7442’, ‘Telaprevir 24 Week+Peg-IFN-alfa-2a 24 Week’, ‘Test Product’, ‘TOTAL 2-sites, TRC’, ‘Cohort 1’, ‘Arm A: hIVIG’, ‘Posaconazole 100 mg QD for 24 Weeks’, ‘Remdesivir’, ‘Menjugate Comparator [6-12W] Group’, ‘HIV-/Cervarix Group’, ‘Vaniprevir 12 Week Arm’, ‘Participants Who Received DTG/3TC FDC’, ‘4-sites, 1-week WITH HRIG’, ‘Daclatasvir, 60 mg, 12-Week Cohort’, ‘Intervention’, ‘13vPnC’, ‘Arm B: ZDV+ABC+3TC+NNRTI->ABC+3TC+NNRTI Maintenance’, ‘VeroRab Comparator [5-17M] Group’, ‘Group D’, ‘PEG 1.0 mcg/kg Weekly (QW) * 24 Weeks’, ‘Cohort 3 eTIV (3-8 Years)’, ‘Sotrovimab (500 mg IV)’, ‘Filgotinib’, ‘rMenB All’, ‘Azelaic Acid Foam, 15% (BAY39-6251)’, ‘QIV-HD’, ‘Placebo’, ‘TIV (3-8 Years)’, ‘Phase 2: 2800 mg Bamlanivimab + 2800 mg Etesevimab’, ‘TDV Lot 2’, ‘13vPnC After Toddler Dose’, ‘AZD1222’, ‘Telaprevir 12 Week+Peg-IFN-alfa-2a, RBV 24/48 Week’, ‘Agriflu’, ‘Period 2: Tacrolimus’]*

### Mesh terms set

*[’taurolidine’, ‘peginterferon alfa-2b’, ‘metronidazole’, ‘chloroquine’, ‘paclitaxel’, ‘acetaminophen’, ‘valganciclovir’, ‘lamivudine’, ‘efavirenz, emtricitabine, tenofovir disoproxil fumarate drug combination’, ‘terbinafine’, ‘loratadine’, ‘vincristine’, ‘adapalene’, ‘sulfadoxine’, ‘everolimus’, ‘antibodies’, ‘interferon alpha-2’, ‘ribavirin’, ‘trimethoprim, sulfamethoxazole drug combination’, ‘rho(d) immune globulin’, ‘immunoglobulins’, ‘methylprednisolone hemisuccinate’, ‘thrombin’, ‘peginterferon alfa-2a’, ‘darunavir’, ‘mycophenolic acid’, ‘alemtuzumab’, ‘diphenhydramine’, ‘nevirapine’, ‘tenofovir’, ‘cyclosporine’, ‘infliximab’, ‘cetuximab’, ‘cyclosporins’, ‘sirolimus’, ‘cobicistat’, ‘promethazine’, ‘gamma-globulins’, ‘trimethoprim’, ‘mometasone furoate’, ‘tofacitinib’, ‘emtricitabine, tenofovir disoproxil fumarate drug combination’, ‘thymoglobulin’, ‘maribavir’, ‘carboplatin’, ‘bamlanivimab’, ‘basiliximab’, ‘pharmaceutical solutions’, ‘gelatin sponge, absorbable’, ‘heparin’, ‘atazanavir sulfate’, ‘azelaic acid’, ‘fanasil, pyrimethamine drug combination’, ‘calcium heparin’, ‘clindamycin’, ‘tazarotene’, ‘vaccines’, ‘posaconazole’, ‘interferons’, ‘olopatadine hydrochloride’, ‘heptavalent pneumococcal conjugate vaccine’, ‘ceftriaxone’, ‘ivermectin’, ‘simeprevir’, ‘rilpivirine’, ‘clotrimazole’, ‘clindamycin phosphate’, ‘ganciclovir’, ‘sulfamethoxazole’, ‘oseltamivir’, ‘abacavir’, ‘raltegravir potassium’, ‘miconazole’, ‘mitogens’, ‘adalimumab’, ‘amoxicillin’, ‘tacrolimus’, ‘nicotinic acids’, ‘antibodies, monoclonal’, ‘epinephrine’, ‘interferon-alpha’, ‘remdesivir’, ‘pyrimethamine’, ‘cobicistat mixture with darunavir’, ‘camostat’, ‘ziprasidone’, ‘efavirenz’, ‘immunoglobulins, intravenous’, ‘artemether, lumefantrine drug combination’, ‘adefovir dipivoxil’, ‘gabexate’, ‘clindamycin palmitate’, ‘letermovir’, ‘methylprednisolone’, ‘ganciclovir triphosphate’, ‘emtricitabine tenofovir alafenamide’, ‘ophthalmic solutions’, ‘zidovudine’, ‘lamivudine, zidovudine drug combination’, ‘prednisone’, ‘emtricitabine’, ‘amodiaquine’, ‘benzoyl peroxide’, ‘azithromycin’, ‘etoposide’, ‘antilymphocyte serum’, ‘bleomycin’, ‘dideoxynucleosides’]*

## Code Availability

The source codes of data collection, processing and analysis are stored at: (https://github.com/xuanyshi/clinical-trials-finer-grained-representation).

## Data Availability

All data produced are available online at https://github.com/xuanyshi/clinical-trials-finer-grained-representation.

## Acknowledgements

This work is funded by the National Key R&D Program for Young Scientists (2022YFF0712000).

We thank Na He (pharmacist, Department of Pharmacy, Peking University Third Hospital) for drug efficacy and adverse events scenarios. We also thank research interns Xiaofan Li (Beijing Institute of Technology), Zitao Liang (Peking University), and Zhanyuan Jiang (Beijing University of Posts and Telecommunications) for help with data processing and analysis.

## Author contributions

Xuanyu Shi was responsible for major data collection, pre-processing and analysis, XS was also responsible for manuscript writing.

Jian Du and Xuanyu Shi conceptualized the research. JD was also responsible for background drafting and manuscript polishing.

---

**Supplementary Table 1.**
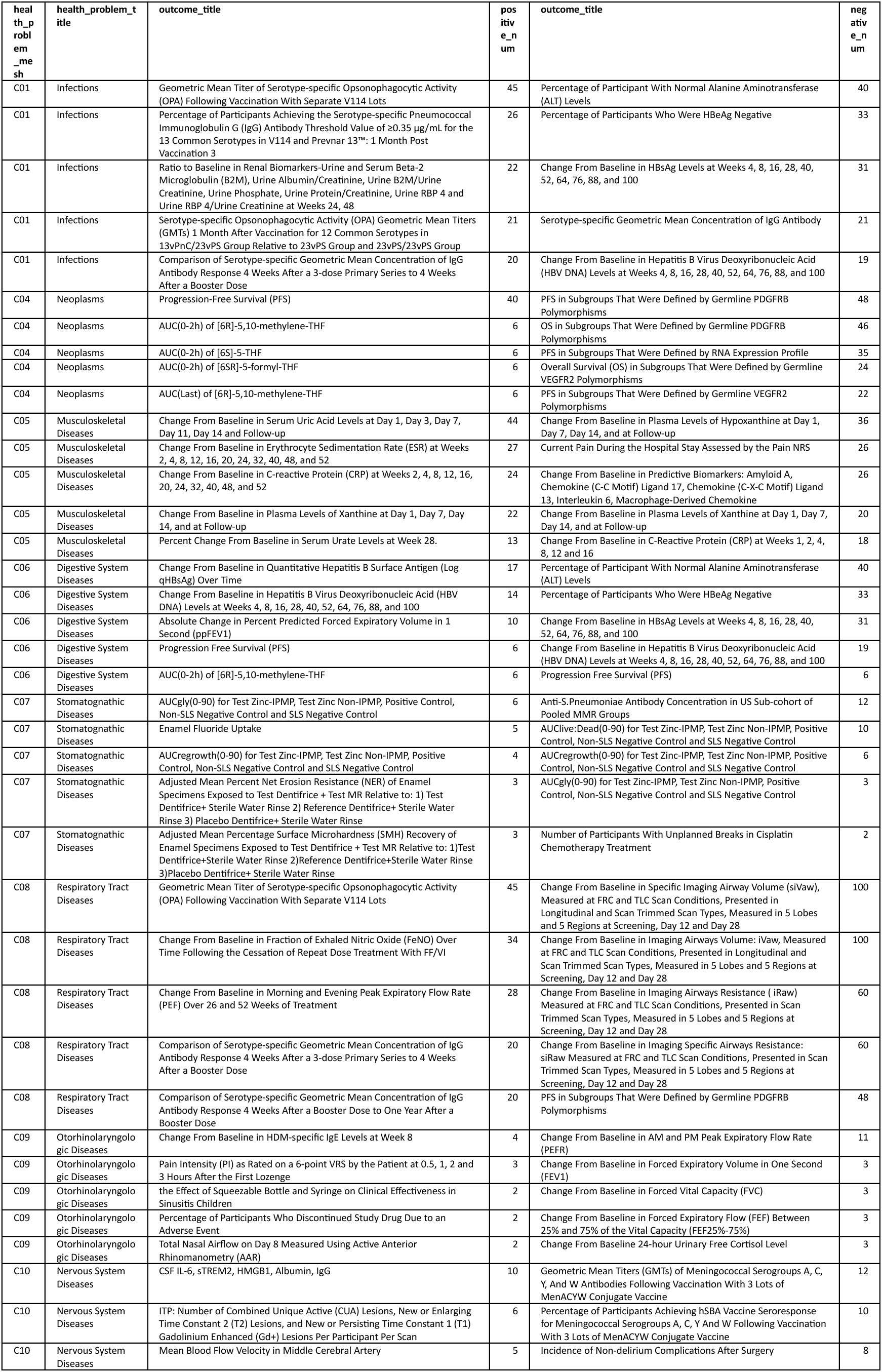
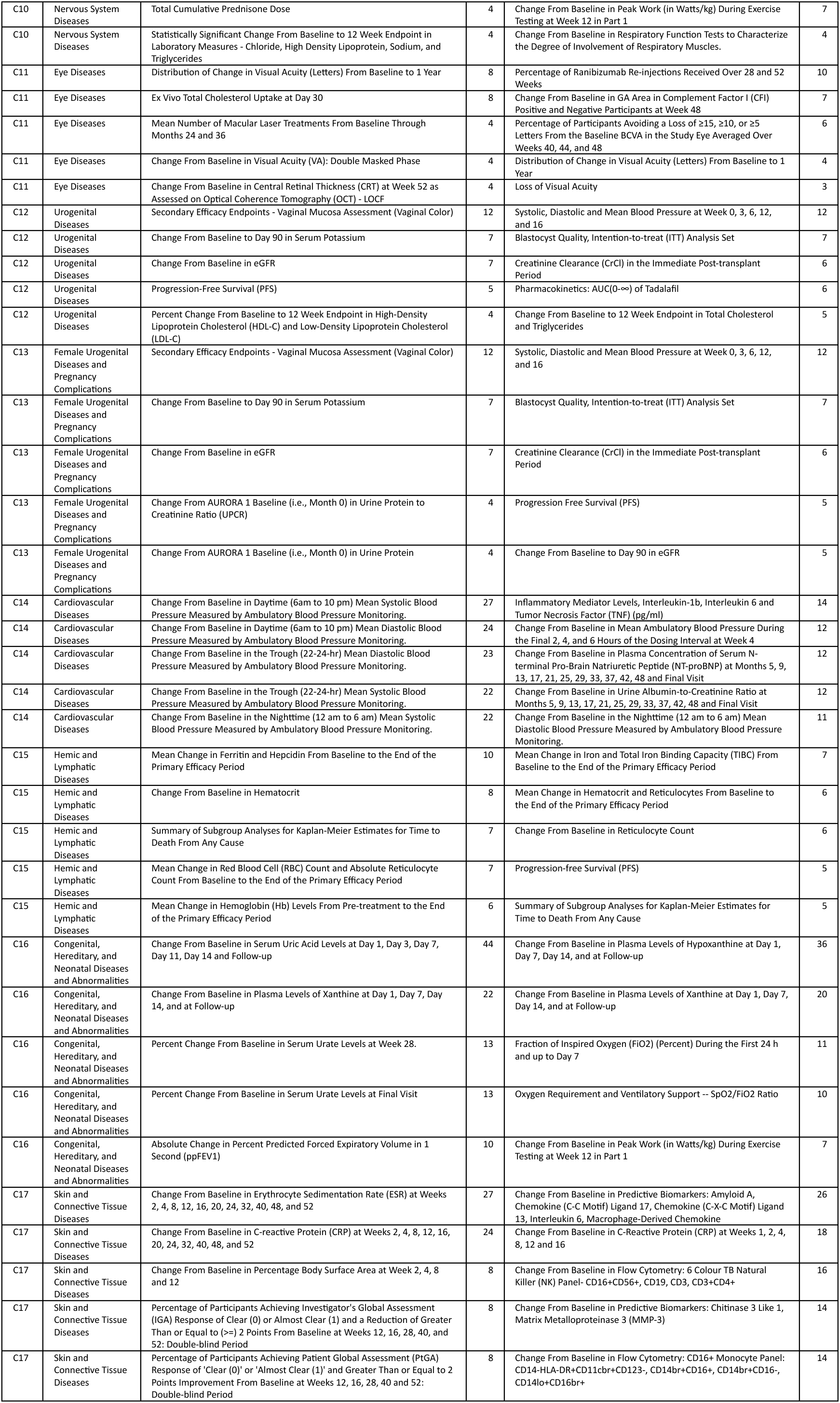
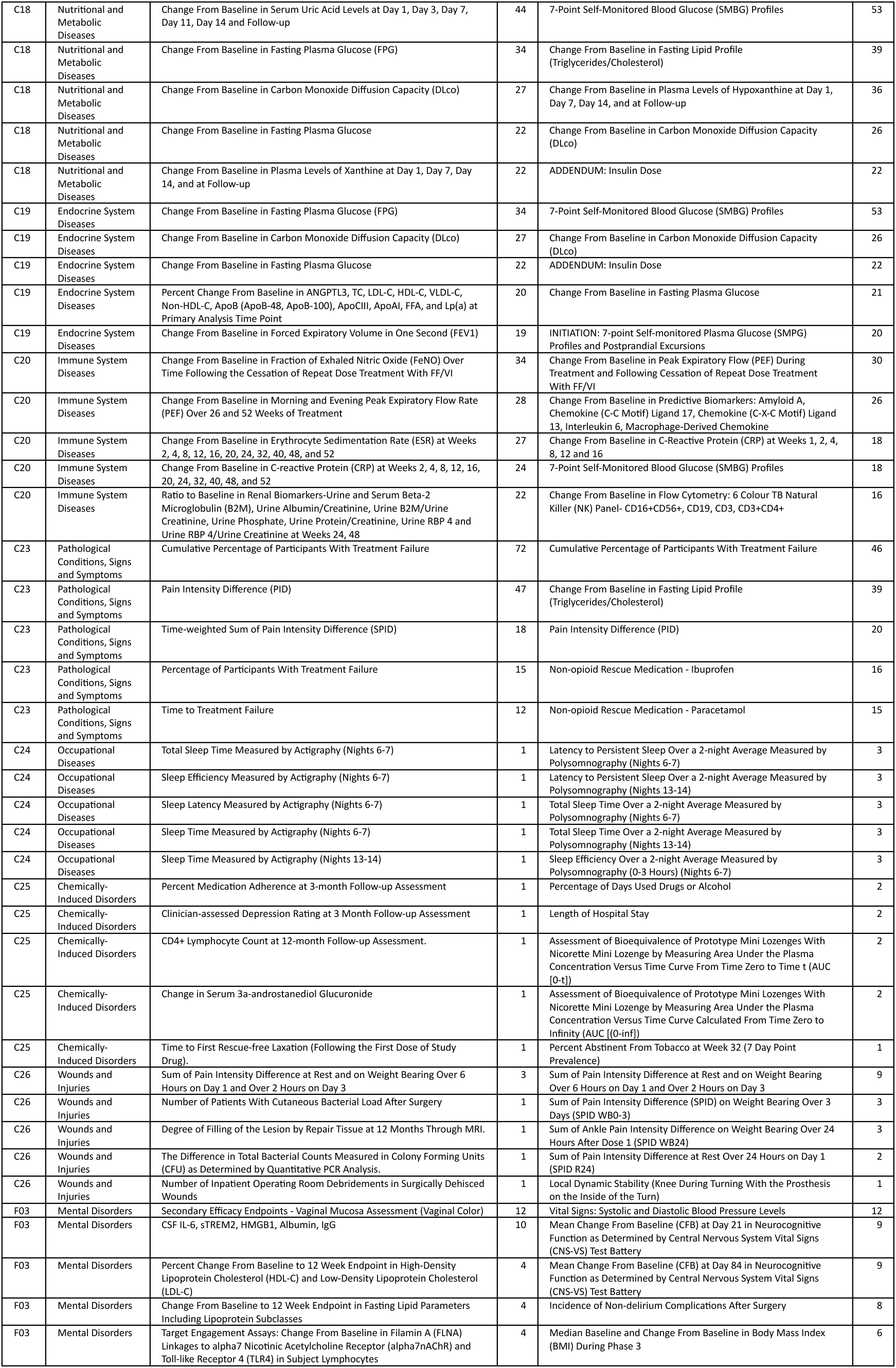
Positive and negative biomarker outcome set of each health problem.

